# Elevated suicidal thoughts and behaviors, proximal suicide risk factors, and non-suicidal self-injury among adults reporting symptoms of Cannabinoid Hyperemesis Syndrome: Results from a national survey of US adults

**DOI:** 10.64898/2026.02.26.26347185

**Authors:** Brian M. Hicks, Blair J. Whittington, Amanda M. Price, Paula Goldman, Mark A. Ilgen

## Abstract

**Background:** Cannabinoid hyperemesis syndrome (CHS), a disorder characterized by severe nausea, vomiting, and abdominal pain among heavy cannabis users. We previously showed that CHS symptoms are associated with several behavioral and psychological characteristics linked to psychosocial impairment. We examined links between CHS symptoms and suicidal thoughts, behaviors, and proximal suicide risk factors.

**Methods:** We used data from the National Firearms, Alcohol, Cannabis, and Suicide survey, a nationally representative survey of 7034 US adults. Items assessed symptoms of CHS and suicidal thoughts and behaviors, proximal suicide risk factors, and non-suicidal self-injury. Comparisons focused on: those with daily cannabis use and CHS symptoms (*n* = 191), those with daily cannabis use without CHS symptoms (*n* = 882), those with past year cannabis use but not daily use (*n* = 1288), and those without past year cannabis use (*n* = 4673).

**Results:** Those with CHS symptoms reported the highest lifetime and past 12 months prevalence of suicidal thoughts and behaviors with nearly all being significantly higher than those with daily cannabis use without CHS symptoms. Those with CHS symptoms also reported higher mean-levels of proximal risk factors for suicide (i.e., perceived burdensomeness, thwarted belongingness, defeat, entrapment) than all the other groups.

**Conclusions:** Those with CHS symptoms reported especially high rates of suicidal thoughts, behaviors, and attempts even when compared to others with daily cannabis use. People with CHS symptoms appear to be at high risk of suicide, possibly related to distress from their gastrointestinal symptoms and psychiatric, substance use, and medical comorbidities.

## Introduction

Cannabinoid hyperemesis syndrome (CHS) is an emerging disorder seen among those who use cannabis frequently over an extended period and experience repeated nausea during the prodromal phase and recurrent episodes of severe nausea, vomiting, and abdominal pain during the acute phase (Allen et al., 2004; Angulo, 2024; Hasler et al., 2025; Jiménez-Castillo et al., 2025; Loganathan et al., 2024; Sorensen et al., 2017; Stjepanović et al., 2025). As cannabis legalization has expanded and contributed to greater access and higher rates of daily use, the increased availability of high potency cannabis products coupled with declining perceptions of risk (ElSohly et al., 2016; Hall & Lynskey, 2020; Spindle et al., 2019) may be contributing to the increasing prevalence of CHS.

Research on CHS is still in a nascent state, but this condition has been described with increasing frequency in acute care settings (Garcia & Rodríguez, 2025; Jack et al., 2025; Meltzer et al., 2025). Using data from the National Firearms, Alcohol, Cannabis, and Suicide (NFACS) survey, we found that the prevalence of CHS symptoms may be higher than previously thought, as nearly 18% of those who reported a period of daily cannabis use in the past 5 years also endorsed episodes of severe nausea, vomiting, and abdominal pain that are consistent with symptoms of CHS, which projects to over seven million US adults (Ilgen, 2026). Those with symptoms of CHS tended to use cannabis heavily and report more symptoms of cannabis use disorder when compared to those who used daily but did not report CHS symptoms (Ilgen, 2026).

Using data from this same survey, those with CHS symptoms also had high rates of use of various drug classes and histories of drug overdoses and alcohol poisoning, alcohol use problems, antisocial behavior, intimate partner violence, and disinhibited personality traits (e.g., antagonism, low conscientiousness, negative urgency) (Hicks et al., 2026). CHS symptoms were also associated with greater depression and anxiety and lower social support and subjective well-being. People with CHS symptoms also reported more health-related problems including more sleep problems, chronic pain, and worse overall health. While those with CHS symptoms had much higher rates of these behavioral and psychological problems than those with low levels of cannabis use, they also reported significantly more of these problems than daily cannabis users without CHS symptoms. These results indicate that those with CHS symptoms carry a high disease burden of comorbid psychiatric, substance use, and medical disorders.

Many of these psychological and behavioral problems have been independently linked to suicidal thoughts and behaviors (Franklin et al., 2017). Given the consistent association between CHS symptoms and these problems, it is reasonable to hypothesize that those with CHS symptoms may be at elevated risk for suicidal thoughts and behaviors. Similarly, heavy cannabis use has been shown to be associated with higher rates of suicidal thoughts and behaviors, with stronger effects seen in younger populations and those with cannabis use disorder (Bohnert et al., 2017; Carvalho et al., 2022; Delforterie et al., 2015; Denissoff et al., 2022; Diep et al., 2022; Fresán et al., 2022). As those with CHS symptoms are more likely to have problem use and other behavioral and psychological risk factors, even relative to other adults who use cannabis daily, it is likely that they represent an extreme end of the continuum of risks associated with heavy cannabis use and may be especially likely to report suicidal thoughts and behaviors.

Conceptualizing suicidal thoughts and behaviors as a continuum of severity, it is also plausible that those with CHS symptoms will report elevated levels of related problems including non-suicidal self-injury and thoughts and feelings hypothesized to be proximal risk factors for suicide. While non-suicidal self-injury has distinctive features from suicidal thoughts and behaviors, there is also substantial overlap in their risk factors and correlates (Anestis et al., 2013; Franklin et al., 2017; Hamza et al., 2012; Klonsky et al., 2016; Muehlenkamp, 2005; Nock & Favazza, 2009; Nock et al., 2006). Cognitive-emotional constructs including perceived burdensomeness (belief that one is a and thwarted belongingness as well as defeat and entrapment which are that are components of the influential Interpersonal Theory of Suicide (Joiner et al., 2009; Van Orden et al., 2010) and the Integrated Motivational-Volitional model of suicide (O’Connor & Kirtley, 2018), respectively, have been extensively studied and conceptualized as proximal risk factors and important for understanding a person’s suicide risk profile and may help explain the association between cannabis use and suicide risk (Baer et al., 2022; Chu et al., 2017). Adding measures of non-suicidal self-injury and proximal suicide risk factors to the assessment of suicidal thoughts and behaviors and covering multiple time frames (e.g., lifetime, past 12 months, past 2 weeks) helps to provide a more comprehensive assessment of suicide-related risk including its chronicity and acuity.

We used the NFACS survey sample, large nationally representative sample of US adults, to examine the associations between CHS symptoms and suicidal thoughts, behaviors, prior attempts, non-suicidal self-injury, and other proximal suicide risk factors. Comparisons between those reporting CHS symptoms and three other groups varying levels of cannabis use including those who reported a period of daily or near daily cannabis use in the past 5 years without CHS symptoms, those with less frequent cannabis use in the past 12 months, or those who reported no cannabis use within the past 12 months. Our goal was to determine if individuals endorsing CHS symptoms are a distinctive group at higher risk for suicide even when compared to others who use cannabis heavily, which may indicate that those with CHS symptoms are an especially high-risk psychiatric group warranting unique targeted suicide prevention and intervention efforts.

## Methods

### Participants

Data were collected as part of the National Firearms, Alcohol, Cannabis, and Suicide survey (*N* = 7,034), a nationally representative survey of U.S. adults aged 18 years or older, the details of which have been described elsewhere (Ilgen, 2026). The survey was developed by the study team and fielded by a market research company, Verasight, from May 27 – September 2, 2025. Participants were recruited via random address-based sampling (ABS) or multimedia/single messaging service (MMS/SMS) methods (aka, text messaging). For the ABS sample, potential participants were identified through the US Postal Service database and mailed an initial invitation letter as well as a reminder postcard. Where corresponding cell phone numbers and email addresses were available, the research company also sent follow-up reminders with a direct URL to the web-based survey utilizing those alternative contact methods. For the MMS/SMS text message recruitment, potential participants were identified from a database consisting of voter file and supplemental commercial records and received a survey link via text message. Participants received payment for participation. Response rates were 3.83% (*n* = 1,914) and 0.39% (*n =* 8,652) for the ABS and MMS/SMS text messaging, respectively. From these responses, 2,829 (26.7%) were removed due to incomplete data. An additional 703 responses (6.7%) were excluded during the quality assurance process, which included confirming all responses corresponded with an IP address in the US, no duplicate responses were provided, no non-human responses were included and removing participants who completed the survey in less than 30% of the median completion time. This yielded a sample of 1,509 ABS-recruited and 5,525 MMS/SMS-recruited participants. Responses were weighted to match national benchmarks from the July 2025 Current Population Survey for age, sex, race/ethnicity, income, education, political party identification, region, and metropolitan status.

The protocol was reviewed by the University of Michigan IRB and was determined to be exempt. Prior to accessing the survey, participants reviewed information on the survey content, length of the survey and compensation, and were informed that participation was voluntary and anonymous. All participants provided active consent prior to receiving survey questions and all participants completed the survey online.

### Assessment

*CHS symptoms.* We asked a series of questions to capture a group of participants whose response profile was consistent with CHS. All respondents who endorsed at least 20 lifetime uses of cannabis were asked if they had a period of daily or near daily cannabis use in the past 5 years. Those who endorsed daily use in the past 5 years were asked if they had experienced periods of severe nausea, vomiting, or abdominal pain. Using these responses, we made four mutually exclusive groups for comparisons: the CHS symptom group included respondents who endorsed a period of daily cannabis use in the past 5 years and periods of nausea, vomiting, or abdominal pain (*n* = 191); all other respondents who endorsed a period of daily cannabis use in the past 5 years but not periods of severe nausea, vomiting, or abdominal pain (*n* = 882); all other respondents who reported cannabis use in the past 12 months (*n* = 1288); all other respondents who we refer to as the no cannabis use in the past 12 months group (*n* = 4673).

*Suicidal thoughts and behaviors and non-suicidal self-injury.* We also included several items to assess suicidal thoughts and behaviors including passive thoughts of death, suicidal ideation, suicide planning, firearm suicidal ideation, suicide preparatory behaviors, aborted suicide attempt, interrupted suicide attempt, and suicide attempt for lifetime and past 12 months (see Tables 1 and 2 for item wording). We also assessed non-suicidal self-injury for lifetime and past 12 months. These items were adapted from existing measures of suicide-related outcomes and were selected to capture a wide range of suicidal experiences (Fox et al., 2020; Hoffmire et al., 2022; Millner et al., 2015; Posner et al., 2011). We also calculated a latent trait or continuous score of lifetime suicide thoughts and behaviors by fitting a two-parameter logistic response model to all the relevant items.

**Table 1.**
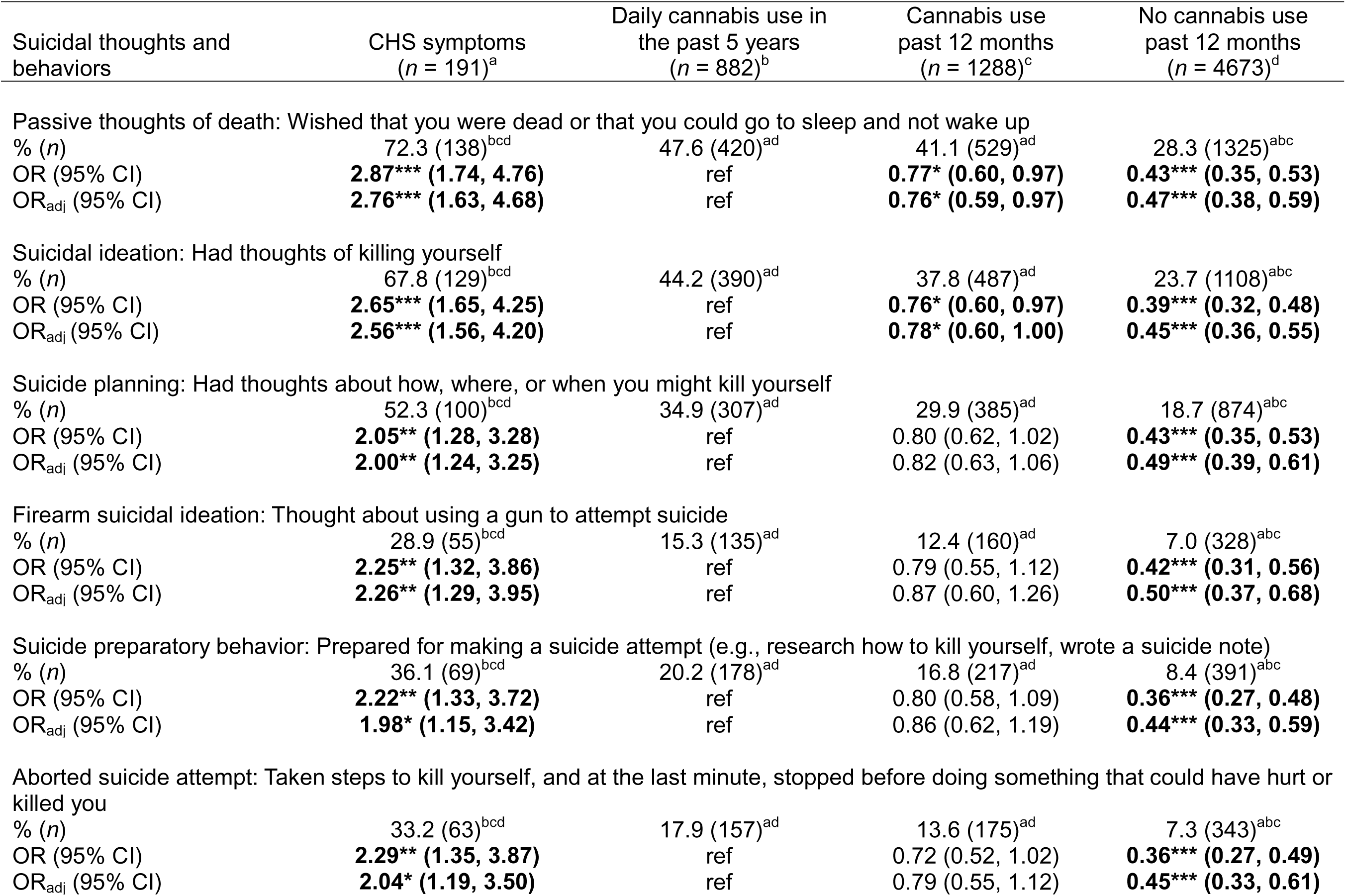
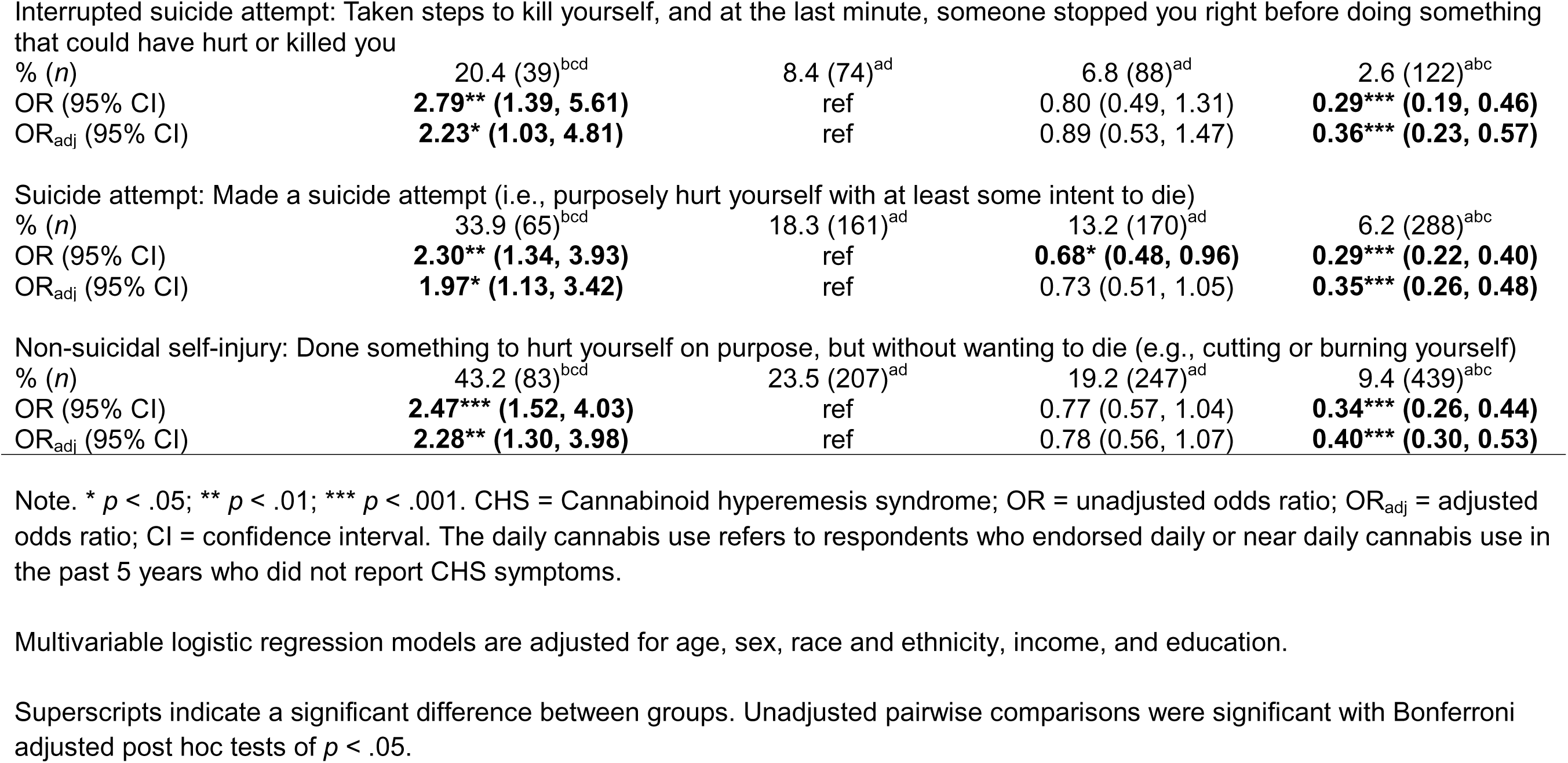
Lifetime suicidal thoughts and behaviors and non-suicidal self-injury among CHS symptom and cannabis use groups; National Firearms, Alcohol, Cannabis, and Suicide survey, 2025 (*N* = 7034)

**Table 2.** Past 12 months suicidal thoughts and behaviors and non-suicidal self-injury among CHS symptom and cannabis use groups; National Firearms, Alcohol, Cannabis, and Suicide survey, 2025 (*N* = 7034)

| Suicidal thoughts and behaviors | CHS symptoms<br>(n = 191)<sup>a</sup> | Daily cannabis use in<br>the past 5 years<br>(n = 882)<sup>b</sup> | Cannabis use<br>past 12 months<br>(n = 1288)<sup>c</sup> | No cannabis use<br>past 12 months<br>(n = 4673)<sup>d</sup> |
| --- | --- | --- | --- | --- |
| Passive thoughts of death: Wished that you were dead or that you could go to sleep and not wake up |  |  |  |  |
| % (n) | 43.5 (83)<sup>bcd</sup> | 20.4 (180)<sup>ad</sup> | 14.6 (188)<sup>ad</sup> | 9.6 (449)<sup>abc</sup> |
| OR (95% CI) | **3.01\*\*\* (1.85, 4.91)** | ref | **0.67\* (0.49, 0.91)** | **0.42\*\*\* (0.32, 0.54)** |
| OR<sub>adj</sub> (95% CI) | **2.72\*\*\* (1.67, 4.44)** | ref | **0.71\* (0.51, 0.99)** | **0.50\*\*\* (0.38, 0.66)** |
| Suicidal ideation: Had thoughts of killing yourself |  |  |  |  |
| % (n) | 29.2 (56)<sup>bcd</sup> | 14.8 (131)<sup>ad</sup> | 11.4 (147)<sup>ad</sup> | 5.7 (265)<sup>abc</sup> |
| OR (95% CI) | **2.37\*\* (1.37, 4.09)** | ref | 0.74 (0.51, 1.07) | **0.35\*\*\* (0.25, 0.48)** |
| OR<sub>adj</sub> (95% CI) | **2.15\*\* (1.22, 3.79)** | ref | 0.80 (0.54, 1.17) | **0.42\*\*\* (0.31, 0.58)** |
| Suicide planning: Had thoughts about how, where, or when you might kill yourself |  |  |  |  |
| % (n) | 27.0 (51)<sup>bcd</sup> | 10.8 (96)<sup>ad</sup> | 8.8 (113)<sup>ad</sup> | 4.8 (226)<sup>abc</sup> |
| OR (95% CI) | **3.03\*\*\* (1.73, 5.32)** | ref | 0.79 (0.53, 1.18) | **0.42\*\*\* (0.29, 0.59)** |
| OR<sub>adj</sub> (95% CI) | **2.81\*\*\* (1.57, 5.02)** | ref | 0.87 (0.58, 1.32) | **0.52\*\*\* (0.36, 0.74)** |
| Firearm suicidal ideation: Thought about using a gun to attempt suicide |  |  |  |  |
| % (n) | 13.9 (27)<sup>bcd</sup> | 5.1 (45)<sup>a</sup> | 4.0 (52)<sup>a</sup> | 2.8 (129)<sup>a</sup> |
| OR (95% CI) | **3.01\*\*\* (1.54, 5.91)** | ref | 0.78 (0.44, 1.39) | **0.53\*\* (0.33, 0.86)** |
| OR<sub>adj</sub> (95% CI) | **2.85\*\* (1.41, 5.77)** | ref | 0.91 (0.50, 1.65) | 0.71 (0.43, 1.18) |
| Suicide preparatory behavior: Prepared for making a suicide attempt (e.g., research how to kill yourself, wrote a suicide note) |  |  |  |  |
| % (n) | 11.1 (21)<sup>bcd</sup> | 3.3 (29)<sup>ad</sup> | 3.2 (41)<sup>ad</sup> | 1.1 (50)<sup>abc</sup> |
| OR (95% CI) | **3.67\*\* (1.51, 8.92)** | ref | 0.97 (0.46, 2.03) | **0.32\*\*\* (0.16, 0.63)** |
| OR<sub>adj</sub> (95% CI) | **2.96\* (1.14, 7.69)** | ref | 1.08 (0.50, 2.32) | **0.41\* (0.20, 0.84)** |
| Aborted suicide attempt: Taken steps to kill yourself, and at the last minute, stopped before doing something that could have hurt or killed you |  |  |  |  |
| % (n) | 8.7 (17)<sup>bcd</sup> | 2.1 (19)<sup>a</sup> | 2.0 (25)<sup>a</sup> | 1.1 (50)<sup>a</sup> |
| OR (95% CI) | **4.38\*\* (1.51, 12.68)** | ref | 0.91 (0.37, 2.24) | 0.49 (0.22, 1.09) |
| OR<sub>adj</sub> (95% CI) | **3.79\* (1.25, 11.49)** | ref | 1.10 (0.43, 2.82) | 0.68 (0.30, 1.53) |
| Interrupted suicide attempt: Taken steps to kill yourself, and at the last minute, someone stopped you right before doing something that could have hurt or killed you |  |  |  |  |
| % (n) | 5.6 (11)<sup>d</sup> | 0.7 (6) | 1.4 (18)<sup>d</sup> | 0.2 (12)<sup>ac</sup> |
| OR (95% CI) | **8.09\* (1.48, 44.28)** | ref | 1.95 (0.42, 8.99) | 0.34 (0.08, 1.46) |
| OR<sub>adj</sub> (95% CI) | **6.49\* (1.07, 39.23)** | ref | 2.36 (0.50, 11.22) | 0.43 (0.10, 1.92) |
| Suicide attempt: Made a suicide attempt (i.e., purposely hurt yourself with at least some intent to die) |  |  |  |  |
| % (n) | 7.0 (13)<sup>bd</sup> | 0.9 (8)<sup>a</sup> | 2.1 (27)<sup>d</sup> | 0.4 (19)<sup>ac</sup> |
| OR (95% CI) | **8.43\*\* (1.96, 36.29)** | ref | 2.37 (0.66, 8.52) | 0.46 (0.13, 1.65) |
| OR<sub>adj</sub> (95% CI) | **6.70\* (1.42, 31.70)** | ref | 2.85 (0.76, 10.74) | 0.64 (0.17, 2.37) |
| Non-suicidal self-injury: Done something to hurt yourself on purpose, but without wanting to die (e.g., cutting or burning yourself) |  |  |  |  |
| % (n) | 17.3 (33)<sup>cd</sup> | 7.7 (68)<sup>d</sup> | 5.9 (76)<sup>ad</sup> | 2.9 (137)<sup>abc</sup> |
| OR (95% CI) | **2.51\* (1.24, 5.11)** | ref | 0.75 (0.45, 1.27) | **0.36\*\*\* (0.23, 0.58)** |
| OR<sub>adj</sub> (95% CI) | 2.21 (0.98, 4.98) | ref | 0.79 (0.46, 1.35) | **0.48\*\* (0.30, 0.76)** |

Note. \* $p < .05$ ; \*\* $p < .01$ ; \*\*\* $p < .001$ . CHS = Cannabinoid hyperemesis syndrome; OR = unadjusted odds ratio; OR<sub>adj</sub> = adjusted odds ratio; CI = confidence interval. The daily cannabis use refers to respondents who endorsed daily or near daily cannabis use in the past 5 years who did not report CHS symptoms.
Multivariable logistic regression models are adjusted for age, sex, race and ethnicity, income, and education.
Superscripts indicate a significant difference between groups. Unadjusted pairwise comparisons were significant with Bonferroni adjusted post hoc tests of $p < .05$ .

*Proximal suicide risk factors.* We included markers of thoughts and feelings over the past 2 weeks that have been associated with suicide risk, specifically, defeatism (2 items, α = .44; e.g., “Felt like I have given up.”) and entrapment (2 items, α = .78; e.g., “Thought that I cannot see a way out of my current situation.”) as described in the Integrated Motivational-Volitional Model of suicidal behavior (Gilbert & Allan, 1998; O’Connor & Kirtley, 2018). Seven items from the short version of the Interpersonal Needs Questionnaire (Van Orden et al., 2012) were selected to measure thwarted belongingness (3 items, α = .64; e.g., “Felt disconnected from other people.”) and perceived burdensomeness (4 items, α = .90; e.g., “Thought that people in my life would be happier without me.”) as detailed in the Interpersonal Theory of suicide (Joiner et al., 2009; Van Orden et al., 2010). We also calculated a latent trait score of suicide risk characteristics by fitting a graded response model to all these items (11 items, α = .89).

*Demographic variables.* Respondents reported on various demographic characteristics including age, sex, race and ethnicity (White, Black, Hispanic, Other), annual household income (< $50,000, $50,000 to $100,000, and > $100,000), and education (high school or less, some college, bachelor’s degree, graduate degree).

### Analyses

Logistic and linear regression models were fit to compare the CHS symptom and other cannabis use groups on the suicidal thoughts and behaviors, non-suicidal self-injury, and proximal suicide risk factors. For each model, the suicide related variable were the outcomes. We fit unadjusted model that only included the four-level CHS symptom and cannabis use groups and adjusted models that also included age, sex, race/ethnicity, income, and education. All possible post hoc pairwise were also tested with a Bonferroni adjustment for multiple comparisons and used a protected alpha of *p* < .05 for statistical significance. Our first goal was to determine if the CHS symptom group differed from the daily cannabis use group. We then compared the CHS symptom and daily use groups to the less frequent cannabis use groups to further assess whether group differences were due to cannabis use in general or if there were unique risks associated with CHS symptoms. All analyses were conducted in Stata 19 using the Survey data analysis module with poststratification weights to adjust parameter estimates and standard errors in all models.

## Results

### Lifetime Suicidal Thoughts, Behaviors, and Attempts and Non-suicidal Self-injury

Table 1 presents the lifetime prevalence rates and group differences for eight forms of suicidal thoughts and behaviors including past suicide attempts as well as non-suicidal self-injury. The CHS symptom group had higher lifetime rates of each of the suicidal thoughts and behaviors and non-suicidal self-injury than all other groups including those with daily cannabis use without CHS symptoms (mean unadjusted and adjusted OR = 2.43 and 2.23, respectively). All the pairwise comparisons between the CHS symptom group and the other cannabis use groups were significant after the Bonferroni adjustment for multiple post hoc comparisons. The daily use and non-daily cannabis use in the past 12 months groups all had higher rates of each type of suicidal thought and behavior than the no cannabis use in the past 12 months group, even after adjusting for multiple comparisons. Those with daily use and non-daily cannabis use in the past 12 months groups were relatively similar; the only significant differences were that those with daily use had higher rates of wishing they were dead, thoughts of killing oneself, and past suicide attempt, but none of these pairwise comparisons were significant after adjusting for multiple comparisons. The CHS symptom group had higher continuous suicidal thoughts and behaviors scores than all the other groups with medium effect size differences for the daily use (*d* = 0.57, *p* < .001) and cannabis use in the past 12 months (*d* = 0.71, *p* < .001) groups and a large difference with the no cannabis use in past 12 months group (*d* = 1.04, *p* < .001) (see Table 3).

**Table 3.** Number of suicide attempts, suicidal thoughts and behaviors, and proximal suicide risk factors among CHS symptom and cannabis use groups; National Firearms, Alcohol, Cannabis, and Suicide survey, 2025 (*N* = 7034)

|  | CHS symptoms<br>(n = 191)<sup>a</sup> | Daily cannabis use in<br>the past 5 years<br>(n = 882)<sup>b</sup> | Cannabis use<br>past 12 months<br>(n = 1288)<sup>c</sup> | No cannabis use<br>past 12 months<br>(n = 4673)<sup>d</sup> |
| --- | --- | --- | --- | --- |
| Number of suicide attempts |  |  |  |  |
| M (SD) | 1.19 (1.89)<sup>bcd</sup> | 0.43 (1.07)<sup>ad</sup> | 0.31 (1.01)<sup>ad</sup> | 0.15 (0.73)<sup>abc</sup> |
| $\beta$ (95% CI) | ref | **-0.76\*\* (-1.25, -0.27)** | **-0.88\*\*\* (-1.36, -0.40)** | **-1.04\*\*\* (-1.51, -0.56)** |
| $\beta_{adj}$ (95% CI) | ref | **-0.70\*\* (-1.18, -0.21)** | **-0.79\*\*\* (-1.27, -0.31)** | **-0.92\*\*\* (-1.40, -0.44)** |
| Suicide thoughts and behaviors trait score – lifetime |  |  |  |  |
| M (SD) | 58.6 (9.5)<sup>bcd</sup> | 52.9 (10.4)<sup>ad</sup> | 51.5 (10.9)<sup>ad</sup> | 48.2 (9.2)<sup>abc</sup> |
| $\beta$ (95% CI) | ref | **-5.65\*\*\* (-7.94, -3.36)** | **-7.10\*\*\* (-9.29, -4.90)** | **-10.44\*\*\* (-12.52, -8.35)** |
| $\beta_{adj}$ (95% CI) | ref | **-5.16\*\*\* (-7.42, -2.90)** | **-6.45\*\*\* (-8.64, -4.26)** | **-9.09\*\*\* (-11.19, -6.99)** |
| Defeat |  |  |  |  |
| M (SD) | 57.9 (10.8)<sup>bcd</sup> | 52.0 (10.2)<sup>ad</sup> | 51.3 (10.8)<sup>ad</sup> | 48.9 (9.4)<sup>abc</sup> |
| $\beta$ (95% CI) | ref | **-5.84\*\*\* (-8.24, -3.43)** | **-6.58\*\*\* (-8.90, -4.27)** | **-8.91\*\*\* (-11.12, -6.69)** |
| $\beta_{adj}$ (95% CI) | ref | **-4.32\*\*\* (-6.50, -2.14)** | **-4.00\*\*\* (-6.10, -1.91)** | **-5.52\*\*\* (-7.54, -3.50)** |
| Entrapment |  |  |  |  |
| M (SD) | 58.8 (11.9)<sup>bcd</sup> | 52.4 (11.1)<sup>ad</sup> | 51.7 (11.4)<sup>ad</sup> | 48.7 (8.9)<sup>abc</sup> |
| $\beta$ (95% CI) | ref | **-6.41\*\*\* (-9.26, -3.55)** | **-7.10\*\*\* (-9.83, -4.37)** | **-10.10\*\*\* (-12.73, -7.48)** |
| $\beta_{adj}$ (95% CI) | ref | **-5.51\*\*\* (-8.38, -2.64)** | **-5.68\*\*\* (-8.43, -2.94)** | **-8.13\*\*\* (-10.80, -5.46)** |
| Perceived burdensomeness |  |  |  |  |
| M (SD) | 60.2 (15.8)<sup>bcd</sup> | 52.2 (11.3)<sup>ad</sup> | 51.2 (12.1)<sup>ad</sup> | 48.8 (8.3)<sup>abc</sup> |
| $\beta$ (95% CI) | ref | **-8.03\*\*\* (-12.19, -3.88)** | **-8.99\*\*\* (-13.07, -4.92)** | **-11.35\*\*\* (-15.33, -7.38)** |
| $\beta_{adj}$ (95% CI) | ref | **-6.98\*\*\* (-11.06, -2.91)** | **-7.17\*\*\* (-11.17, -3.18)** | **-9.08\*\*\* (-13.01, -5.15)** |
| Thwarted belongingness |  |  |  |  |
| M (SD) | 58.2 (12.0)<sup>bcd</sup> | 51.9 (10.6)<sup>ad</sup> | 51.4 (10.7)<sup>ad</sup> | 48.9 (9.3)<sup>abc</sup> |
| $\beta$ (95% CI) | ref | **-6.26\*\*\* (-9.12, -3.40)** | **-6.81\*\*\* (-9.57, -4.06)** | **-9.27\*\*\* (-11.95, -6.59)** |
| $\beta_{adj}$ (95% CI) | ref | **-4.95\*\*\* (-7.62, -2.29)** | **-4.52\*\*\* (-7.07, -1.97)** | **-6.41\*\*\* (-8.90, -3.92)** |
| Suicide risk traits |  |  |  |  |
| M (SD) | 60.0 (9.1)<sup>bcd</sup> | 53.6 (10.1)<sup>ad</sup> | 52.5 (10.7)<sup>ad</sup> | 49.1 (9.7)<sup>abc</sup> |
| $\beta$ (95% CI) | ref | **-6.46\*\*\* (-8.55, -4.38)** | **-7.59\*\*\* (-9.56, -5.61)** | **-10.96\*\*\* (-12.83, -9.09)** |
| $\beta_{adj}$ (95% CI) | ref | **-5.11\*\*\* (-7.08, -3.13)** | **-5.36\*\*\* (-7.22, -3.49)** | **-7.93\*\*\* (-9.72, -6.13)** |

Note. CHS = Cannabinoid hyperemesis syndrome; M = Mean; SD = Standard deviation; $\beta$ = unadjusted regression coefficient; $\beta_{adj}$ = adjusted regression coefficient; CI = confidence interval.
Multivariable linear regression models are adjusted for age, sex, race and ethnicity, income, and education.
Superscripts indicate a significant difference between groups. Unadjusted pairwise comparisons were significant with Bonferroni adjusted post hoc tests of $p < .05$ .

### Past 12 months Suicidal Thoughts, Behaviors, and Attempts

Table 2 presents the past 12-month prevalence rates and group differences for the eight suicidal thoughts and behaviors including past suicide attempts as well as non-suicidal self-injury. The CHS symptom group had higher past 12 months rates of each type of suicidal thought and behavior and non-suicidal self-injury than all other groups including those with daily cannabis use without CHS symptoms (mean unadjusted and adjusted OR = 4.28 and 3.63, respectively). Seven of the nine pairwise comparisons with the daily cannabis use and non-daily cannabis use in the past 12 months groups were significant after the Bonferroni adjustment for multiple comparisons, while all the pairwise comparisons with the no cannabis use in the past 12 months were significant. The lack of statistical significance for some of the pairwise comparisons between the CHS symptom group and other cannabis use groups was likely due to the low prevalence of suicidal thoughts and behaviors in the past 12 months as the effect sizes were larger than those for the comparisons for lifetime prevalence.

The daily cannabis group reported higher rates of passive thoughts of death in the past 12 months than the cannabis use in the past 12 months group, but this difference was not significant after adjusting for multiple comparisons nor were the differences for any of the other suicidal thoughts and behaviors. The daily cannabis use and non-daily cannabis use in the past 12 months groups had higher rates of passive thoughts of death, suicidal thoughts and planning, preparatory behaviors for a suicide attempt, and non-suicidal self-injury in the past 12 months than the no cannabis use in the past 12 months group. The cannabis use in the past 12 months group also reported higher rates of making a suicide attempt and being interrupted while making a suicide attempt in the past 12 months than the no cannabis use in the past 12 months group.

### Number of Suicide Attempts and Proximal Suicide Risk Factors

Table 3 presents the group differences for number of past suicide attempts and proximal suicide risk factors (i.e., past two weeks). The CHS symptom group reported a greater mean number of suicide attempts than all the other cannabis use groups (*d*’s ranged from 0.49 to 0.73). The CHS symptom group also reported higher mean-levels of each proximal suicide risk factor than all the other cannabis use groups. Differences with the CHS symptom tended to be of large effect size for comparisons with the no cannabis use in the past 12 months group (mean unadjusted and adjusted *d* = 1.01 and 0.74, respectively) and medium for the daily (mean unadjusted and adjusted *d* = 0.66 and 0.54, respectively) and less frequent cannabis use (mean unadjusted and adjusted *d* = 0.74 and 0.53, respectively) groups. The largest group differences were observed for perceived burdensomeness. None of the comparisons between the daily and less frequent cannabis use groups were significant after adjusting for multiple comparisons, though both exhibited small but significant differences with the no cannabis use in the past 12 months group for each proximal suicide risk factor (mean d = 0.36 and 0.27 for the daily and non-daily cannabis use groups, respectively).

## Discussion

CHS is an emerging disorder among those with heavy cannabis use characterized by episodes of severe nausea, vomiting, and abdominal pain (Angulo, 2024; Hasler et al., 2025; Jiménez-Castillo et al., 2025; Loganathan et al., 2024; Sorensen et al., 2017). Previously, we showed that even compared to adults with daily cannabis use, those with CHS symptoms also had higher rates of cannabis use problems, other drug use, mental health problems, antisocial behavior, dysfunctional personality traits, and other health-related problems (Hicks et al., 2026; Ilgen, 2026). We extended those findings by demonstrating that those with CHS symptoms also have elevated levels of suicidal thoughts and behaviors including prior suicide attempts and current suicide risk factors.

It is important to put the levels of suicidal thoughts and behaviors exhibited by those with CHS symptoms into context. Compared to those with no cannabis use in the past 12 months–similar to a healthy control group–there was, on average, an over 400% increase in the odds (mean OR = 5.26) that a person with CHS symptoms would report lifetime suicidal thoughts and behaviors. However, even when compared to other adults who used cannabis daily who are also at elevated risk of various psychological and behavioral problems, there was, on average, an almost 150% increase in the odds that those with CHS symptoms would report lifetime suicidal thoughts and behaviors. That is, those endorsing symptoms consistent with CHS are at notably higher risk of experiencing suicidal thoughts and behaviors even when compared to those use daily but who do not experience CHS symptoms.

The high rates of suicidal thoughts and behaviors among those with CHS symptoms most likely stems from the combined influence of multiple medical, psychiatric, and substance-related problems. People with CHS symptoms have high rates of drug use that are associated with negative consequences for their physical and psychological health, as well as a variety of risk factors that hinder psychosocial functioning and contribute directly and indirectly to increased suicide risk (Carvalho et al., 2022; Delforterie et al., 2015; Diep et al., 2022; Franklin et al., 2017). The cumulative effect of all these problems and difficulty in finding relief likely leads to thoughts and feelings associated with suicide such as defeat, entrapment, and burdensomeness as well as lower subjective well-being (Blais & Grimm, 2025; Hill & Pettit, 2014; Jacobucci et al., 2023; Wang et al., 2023). The recurrent and seemingly unpredictable cyclical episodes of severe nausea, vomiting, and abdominal pain that characterized CHS symptoms can cause substantial physical discomfort and interference with general functioning. It is likely that this can either cause or exacerbate conditions like depression, anxiety, and other health concerns and lead to greater hopelessness or thoughts of suicide. All these factors accumulate a high-risk loading for suicide, with future longitudinal work needed to understand the unique and combined impact of these factors on subsequent suicidal thoughts and behaviors.

Experiencing suicidal thoughts and behaviors is not the same as dying by suicide, and a goal of future research will be to determine if those with CHS have higher rates of suicide mortality relative to adults with daily and less frequent cannabis use as their risk profile would suggest. For one, those with CHS symptoms have high rates of opioid and sedative use as well as high rates of alcohol use problems (Hicks et al., 2026). The hazardous use of these drugs especially in combination with alcohol has high overdose potential and risk of death, and those with CHS symptoms report high rates of prior non-fatal overdoses (Hicks et al., 2026). Two, those with CHS symptoms reported higher rates of thoughts of using a firearm to attempt suicide and use of lethal means is associated with a higher likelihood of death associated with an attempt (Cai et al., 2022; Conner et al., 2019). Third, those with CHS symptoms had high rates of suicide attempts, and actual attempts, even with less lethal means, increase the likelihood of completed suicide (Favril et al., 2023).

This study had several limitations. Although the overall sample was large, the low prevalence of many suicidal thoughts and behaviors, especially in the past 12 months, made it difficult to detect significant group differences after adjusting for multiple comparisons. The consistency of the effect sizes across lifetime and past 12 months for suicidal thoughts and behaviors and the past 2 weeks for the suicide risk traits indicates that the degree of association is similar, with the lack of significant differences likely attributable to insufficient statistical power. The CHS items had few details regarding the history, duration, and intensity of symptoms and did not rule out possible alternative conditions to account for the episodes of nausea and vomiting. The cross-sectional design also limits the ability to make inferences about temporal and, therefore, causal nature of the association between CHS symptoms and suicidal thoughts and behaviors. Further, because the survey was a self-administered questionnaire, interviewers trained in the assessment of CHS could not ask follow-up questions or clarify any potential misinterpretations by respondents or make clinical judgements as to whether the responses were consistent with CHS. Finally, there are general limitations associated with the survey design including the need for online access and proficiency in English to complete the survey, and respondents may have misinterpreted certain questions. Also, while the survey weights help to better approximate the target population, unmeasured variables can still influence potential non-response bias.

Mounting evidence indicates that adults reporting CHS symptoms are a unique group with high morbidity across multiple domains that appear to be more than a direct consequence of heavy cannabis use. The cumulative effect of the comorbidities among medical, psychiatric, and substance use disorders seems to reduce the quality of life and may elevate levels of suicidal thoughts and behaviors. This creates substantial challenges for health care providers. Treatment providers should be aware that those with CHS could be experiencing an array of behavioral health needs in addition to the physical symptoms of CHS and may be at elevated risk for suicidal thoughts and behaviors. Thus, comprehensive treatment for individuals who present with CHS symptoms should address a range of medical, substance-related, and mental health treatment needs. More generally it will be important to determine if CHS symptoms per se are the cause of suicidal thoughts and behaviors or another factor that increases risk among a vulnerable population. It will also be important to continue to refine the measurement of CHS, especially to improve the precision of identifying possible or probably cases which might affect associations with suicide risk. As the prevalence of CHS is increasing in the context of greater cannabis legalization and accessibility, it is important for researchers and treatment providers to develop comprehensive strategies to help with CHS symptoms for this growing and complex clinical problem.

## Financial support

Research reported in this publication was supported by the National Institute of Mental Health award RF1MH137443 (Ilgen, Hicks) and K18 MH135466 (Hicks) of the National Institutes of Health, Research Career Scientist award RCS 19-333 (Ilgen) of the Department of Veterans Affairs, and 2025 Pilot award from the University of Michigan Institute for Firearm Injury. The sponsors had no role in the design, collection, management, analysis, or interpretation of data, the writing of the manuscript, or submission for publication. Dr. Hicks and Dr. Ilgen had full access to all the data in the study and take responsibility for the integrity of the data and the accuracy of the data analysis. The content is solely the responsibility of the authors and does not necessarily represent the official views of the National Institutes of Health or the Department of Veterans Affairs.

## Ethical standards

The authors assert that all procedures contributing to this work comply with the ethnical standards of the relevant national and institutional committees on human experimentation and with the Helsinki Declaration of 1975, as revised in 2008.

## Competing interests

The authors declare no competing interests.

## Data Availability

The data will deposited in the National Data Archive maintained by the National Institutes of Health USA.

## Notes

### Competing Interest Statement

The authors have declared no competing interest.

### Funding Statement

Research reported in this manuscripot was supported by the National Institute of Mental Health award RF1MH137443 (Ilgen, Hicks) and K18 MH135466 (Hicks) of the National Institutes of Health, Research Career Scientist award RCS 19-333 (Ilgen) of the Department of Veterans Affairs, and 2025 Pilot award from the University of Michigan Institute for Firearm Injury. The sponsors had no role in the design, collection, management, analysis, or interpretation of data, the writing of the manuscript, or submission for publication. Dr. Hicks and Dr. Ilgen had full access to all the data in the study and take responsibility for the integrity of the data and the accuracy of the data analysis. The content is solely the responsibility of the authors and does not necessarily represent the official views of the National Institutes of Health or the Department of Veterans Affairs.

### Author Declarations

The protocol was reviewed by the University of Michigan Health Sciences and Behavioral Sciences (HSBS) IRB and received an exempt determination.

### Summary of Updates

This version of the manuscript added analyses and updated information about methods.

